# What constitutes ‘poor’ adherence to medical advice for chronic diseases? Insights from a qualitative study among hypertension and diabetes patients in urban informal settlements, Mumbai Metropolitan Region

**DOI:** 10.1101/2025.05.07.25326926

**Authors:** Jennifer Spencer, Manjula Bahuguna, Sudha Ramani, Sweety Pathak, Sushma Shende, Shanti Pantvaidya, Vanessa D’Souza, Anuja Jayaraman

## Abstract

**Introduction:** The problem of poor adherence to medical advice in the case of non-communicable diseases, the reasons thereof, and how these are exacerbated in low- and middle-income countries (LMICs) is well-recognized. However, there is less clarity on what ‘poor’ adherence encompasses in these settings. Conventional classifications treat ‘poor’ adherence as a singular category, often disregarding its multifaceted nature. Through this study, we aimed to explore the nuances of what constitutes ‘poor’ adherence to medical advice for chronic diseases in vulnerable LMIC settings. This was done by examining the different ways in which hypertension and diabetes patients living in urban informal settlements in the Mumbai Metropolitan Region attempted to adhere to medical advice.

**Methods:** This is a descriptive qualitative study based on in-depth interviews with 26 hypertension and diabetes patients residing in three urban informal settlements in the Mumbai Metropolitan Region. Emerging patterns of adherence were coded and categorized using grounded analysis.

**Findings:** The study shows complex adherence patterns of patients who navigated the lived realities of urban informal settlements while attempting to adhere to medical advice. Hypertensive and diabetic patients in our setting adopted multiple ways of following medical advice related to medication, lifestyle changes and follow-ups. For instance, in attempting to adhere to prescribed medication, patients adjusted or skipped doses, mixed allopathic and alternative medication, switched medication without consultation with doctors, took breaks, or stopped their medications altogether. Patients also adhered well to some aspects of their medical advice and not to others. For instance, some followed dietary recommendations but did not adhere to medication advice.

**Conclusions:** By understanding the nuances and complexities of ‘poor’ adherence in urban informal settlements, the study builds an empirically grounded typology on adherence. Such a typology is useful for research and practice on improving adherence to medical advice in vulnerable LMIC settings.

## 1. Introduction

Globally, it is well recognized that poor adherence to medical advice for chronic diseases can have serious health repercussions, contributing to an increase in the severity of symptoms, complications, and risk of mortality [1–3]. Tackling this issue is crucial for the success of global and national commitments to control chronic conditions [4]. Several studies have highlighted the problem of poor adherence to medical advice in the case of chronic conditions. These studies discuss multiple reasons for poor adherence, including low awareness, a lack of trust in health providers, and financial constraints arising from poverty [5–7] and emphasize how these are exacerbated in resource-constrained contexts of Low- and Middle-Income Countries (LMICs) [8,9]. However, while studies usually discuss reasons for poor adherence, the subtleties of what constitutes ‘poor’ adherence needs more clarity. At present, there is less literature that is drawn from the empirical experiences of patients from vulnerable spaces in LMICs on what adherence encompasses as a conceptual entity in these settings.

This study aimed to add conceptual clarity to the nuances of ‘poor’ adherence from empirical research that is based in urban informal settlements in the Mumbai Metropolitan Region, India. This study qualitatively explores what constitutes ‘poor’ adherence to medical advice by tracing the ways in which hypertension and diabetes patients living in these settlements attempt to adhere to medical advice.

Among chronic diseases in India, there is a high prevalence of hypertension and diabetes at 35.5% and 11.4%, respectively, with a higher burden in urban areas compared to rural areas [10]. We, therefore, chose to study the two chronic diseases of hypertension and diabetes to contribute to a deeper understanding of the phenomena of adherence in the urban informal settlement contexts of LMICs. An increasing burden of non-communicable diseases (NCDs) has been noted in such spaces [11–13]. At the same time, there is evidence of suboptimal disease management practices in these settings across LMICs [14–17]. Improved understanding of poor adherence to medical advice from these settings can enable us to frame contextualized intervention strategies better suited to patients’ needs in urban informal settlements.

### Existing conceptualizations of adherence

Adherence has for long been defined as the compliance to treatment as prescribed by the healthcare provider [3,18]. More recently, such an approach has been criticized as coercive and compelling towards patients [19]. Recognizing the patients’ agency in their treatments, a patient-centered approach was consequently formulated as concordance between the patient and health care provider-where the treatment is decided by consulting the patient, with full knowledge and awareness of the patient and their agreement to the nature of treatment offered [18,19].

Based on these definitions, poor adherence has been understood as any deviance from compliance to treatment or lack of concordance in treatment decisions between patients and healthcare providers [19]. The categorizations of poor adherence have most commonly been understood as unintentional (beyond patients’ choice or control) or intentional (personal motivations/choice of patients) [18–21]. Some authors categorize poor adherence temporally, based on the lifecycle of treatment, such as primary (during treatment initiation) and secondary (after treatment initiation) [22] or as poor adherence in initiation, implementation, and discontinuation of treatment [20].

Some studies are also found to use researcher-determined criteria of non-adherence or poor adherence, which are measured quantitatively. For instance, poor adherence is measured as the number of days in a week when patients skipped medication [11]. Various authors have focused on measuring adherence through quantitative means using scores and scales such as the Morisky Medication Adherence Scale MMAS-8 and Medication Adherence Report Scale MARS-5. These scales use structured surveys, most commonly measuring forgetfulness and breaks in medicines [23–25].

While the definitions of adherence have evolved over time, moving from being coercive to more patient-centric, the classifications of adherence so far have placed less emphasis on the different, nuanced ways in which people engage with medical advice. Further, the phenomenon has not been widely explored qualitatively from a patient perspective in LMICs-despite a few exceptions [6,26]. Our qualitative inquiry conceptualizes adherence based on patient experiences in urban informal settlements, thereby offering alternatives to categories otherwise used to frame universal understandings of adherence [27].

## 2. Methods

### 2.1 Setting

The study was conducted in three urban informal settlements of a municipal corporation with a population of 0.7 million in the Mumbai Metropolitan Region, Maharashtra, India. This study was part of larger research conducted to understand care-seeking for hypertension and diabetes in urban informal settlements in the region [28]. Almost half of the population in the municipal corporation lives in informal settlements [29] that have grown around the industries in the region. Many such settlements house migrant populations working in these industries. The study area has access to a mix of public and private healthcare providers. First-contact care was usually sought by people from private, non-allopathic doctors who had small clinics in these settlements. Some patients also visited private allopathic doctors. The public health system, including one secondary-level hospital and 15 primary-level facilities in the area, was not usually accessed for NCD care.

### 2.2 Study design

The study employed qualitative methods to examine patterns of adherence to medical advice. In the study, we iteratively analyzed and coded data to build context-specific theorizations of adherence while also learning from existing concepts and theorizations [30, 31]. The data comprises interviews with patients diagnosed with hypertension or diabetes (or both) who shared stories on ways in which they adhered to medical advice given to them.

### 2.3 Participant selection

This study was conducted in the intervention areas of the Society for Nutrition, Education and Health Action (SNEHA), a non-governmental organization working in urban informal settlements in Mumbai since 1999. The field staff in SNEHA has strong personal ties with the community and initially identified patients based on the diversity criteria suggested by the research team. The field staff works in the area to implement various initiatives by SNEHA, for which they regularly conduct home visits. During these visits, they identified adults (18 years and above) from the family who had hypertension and diabetes. They explained the purpose of the study to patients and inquired about their willingness to participate. Thereafter, researchers from SNEHA conducted the interviews and discussions at the participants’ convenience and after a formal process of informed consent. Patients were purposively sampled based on diversity in age, gender and years of diagnosis/treatment of the disease.

A total of 26 patients were interviewed, of which 18 were above 50 years of age. An equal number of men and women patients (13 each) were interviewed. More than half (17) of the participants had resided in the area for more than twenty years. Twelve participants were currently employed, while the rest were either unemployed or retired. Nine were diagnosed with hypertension, 10 with diabetes and 7 with both hypertension and diabetes. Eleven participants were diagnosed two years prior to the interaction, 9 had been diagnosed between 3 and 10 years prior to the interaction, and 6 were diagnosed more than ten years prior to the interaction.

### 2.4 Data collection

Data collection was done from 30^th^ September to 18^th^ November 2022. A total of 26 in-depth interviews were conducted with hypertension and diabetes patients in either Hindi or Marathi languages. Authors MB, JS, and SR conducted the interviews; all three are well-trained in qualitative data collection methods and familiar with the local context. The interviews with patients were conducted in their homes and lasted for 30 minutes on average. We used an in-depth interview guide for data collection. The interview guide was developed using existing literature in the field [32–34]. This interview guide was used for a larger study aimed at understanding the care-seeking journeys of NCD patients within the informal settlements [28]. The key points discussed during the interviews have been mentioned in Table 1.

**Table 1:**
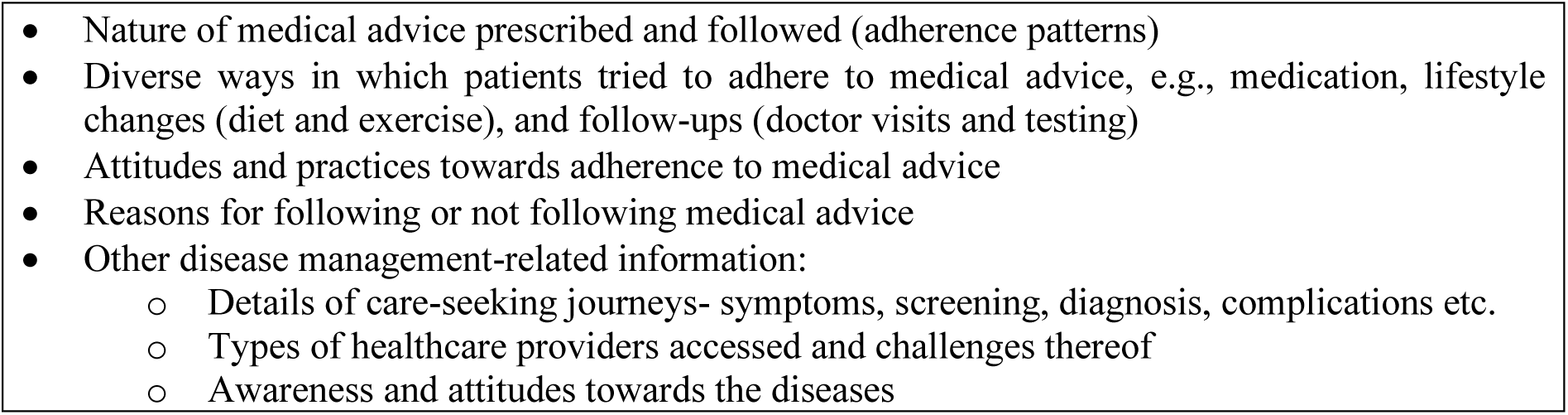
Key points discussed with the participants.

As is the practice in qualitative studies, the exact sample size was not pre-determined [35]. The concept of information redundancy was used as a criterion to ensure data saturation until which the recruitment of additional participants was done [36].

### 2.5 Data analysis

The process of data analysis was done simultaneously with data collection [37]. After every visit, authors JS, SR, MB, and AJ conducted data debriefing sessions to discuss learnings and emerging themes. All interviews were audio-recorded and thereafter simultaneously translated and transcribed into English for analysis. Authors JS, SR, MB and AJ discussed emerging ideas from the transcripts to arrive at a standard set of codes. The transcripts were sorted and coded using NVivo Version 10.3. Substantive coding [38], used in grounded analysis which includes open and axial coding, was done for framing adherence typologies. The various types of adherence as reported by patients were separately coded as initial (open) codes through a line-by-line reading of the transcripts in detail. These were categorized into focused or axial codes consisting of adherence typologies that club several initial codes. We further clustered the emerging patterns of adherence from the data into three broad inductive categories-adherence to prescribed medications, adherence to lifestyle change recommendations, and adherence to instructions for follow-up visits and testing. We have used these three categories to present our findings. Further, we have also showcased case stories and patient narratives under each category to illustrate the nuances of adherence in our setting.

### 2.6 Ethical considerations

Ethical clearance for the research was obtained from ethics committee of the Sigma Research and Consulting Private Limited, India, which gave approval for the study. Recorded verbal informed consent was obtained from the participants. The use of verbal consent was approved by the review committee, owing to COVID-19 guidelines during the data collection period. All the participants gave permission to audio record the interviews. The consent and ensuing interactions were recorded on a smartphone device.

## 3. Findings

The study explores the nuances of ‘poor’ adherence in urban informal settlements using patient narratives on how they attempted to adhere to medical advice. Medical advice has been categorized into medication, lifestyle changes and medical follow-ups.

### 3.1 ‘Poor’ adherence: Categorizing patients’ attempts at adhering to medical advice

#### 3.1.1 Medication

Long-term medication was a primary component of medical advice given to patients. In our setting, most patients interviewed reported being unable to completely adhere to the prescribed medication regimens, though they tried to do so in multiple ways and to varying extents. The different ways they partially adhered to their prescriptions are described as follows.

##### Making4 minor adjustments to medication

Some patients reported making minor adjustments to prescribed medication without consulting their doctors. Patients either adjusted their doses or adjusted the dosage timings. Some reported skipping medicine once in a while. Minor adjustments to medicine were made for many reasons. One reason was forgetting to take medicine. Other reasons included forgetting to replenish medicine strips or being unable to do so on time, one such patient story is elaborated in Case 1.

###### Case 1: Adjusting medicine dosage

A hypertensive patient reported skipping medicines for a few days once a while. This is because he found it difficult to get affordable medicines in the vicinity and did not trust the local hospital to provide medicines for hypertension. During his initial diagnosis, he had visited several private doctors in his locality without success. He had finally been diagnosed and operated on for bypass surgery at a public hospital 50 km away in another city. To date, he prefers to get medicines only from that hospital. Although he tries to take medicines regularly, there is a time gap between his medicines getting over and him going to get a new stock of medicine. Due to this, he either skips medicines for a few days or, if there are fewer tablets left, he reduces the doses to make the strip last longer. – “*If there is less medicine at times, I eat accordingly or manage.*” (Patient, male, diagnosed 6 to 10 years prior to the interaction, age group 46 to 55 years)

This case highlights how unavailability of quality public health services in the vicinity of the urban informal settlement, led patients to make minor adjustments to their medication regimes.

Patients who made such adjustments, like in Case 1, often believed that these did not adversely impact their disease prognosis or health; they looked at these adjustments as minor practical tweaks rather than deviations from prescriptions.

##### Mixing allopathic and alternative medicine

In the vicinity of the urban informal settlements, a range of alternative medicine providers (with degrees or diplomas in Ayurveda, Unani, or Homeopathy) and allopathic providers had their practice. Some patients mentioned alternating or mixing medications prescribed or obtained from both allopathic and alternative medicine practitioners. Patients shared that mixing medications could provide faster relief from symptoms, enabling them to easily get back to their daily work or household activities. Patients also believed alternative medications to have fewer side effects, preferring them for the longer term. It was also shared that alternative medication was cheaper in comparison to allopathy.

Case 2 illustrates the medication regime of one patient who routinely used ayurvedic medicine to alleviate her symptoms and used allopathic medicine only when the symptoms became severe.

###### Case 2: Mixing medications

A patient was diagnosed with both hypertension and diabetes reported taking only diabetes medicines prescribed by a local doctor, which she felt *“did not suit”* her. She went to another private doctor who prescribed new medicines that she felt suited her “*a little*.” After changing multiple doctors, she went to a trust hospital (in another city outside the metropolitan region) whose allopathic medicines she took for a few months.

However, due to financial constraints, she stopped those medicines (Rs. 300 per strip, lasts ten days) and only kept some for emergencies-*“The allopathic medicines benefit me more, but I use them only for emergency purposes.”* Instead, she buys a cheaper ayurvedic syrup (Rs. 200 per bottle, lasts more than a month) from a local ayurvedic vendor-*“If we have money, I get the allopathic medicine. Otherwise, I take ayurvedic medicine.”* According to her, ayurvedic medicine is cheaper but does not provide instant relief. To alleviate her symptoms, once in a while, she purchases quick-fix allopathic medicines such as sleeping pills: *“I do have problems due to not taking (allopathic) medicine. All night, I sit until its morning. I feel pressure in my chest and a feeling of beating hard and fast. If I feel very bad, then I go and get the sleeping pill for Rs. 40 and sleep a little peacefully.”* (Patient, female, diagnosed upto 2 years prior to the interaction, age group 56 to 65 years)

Case 2 also shows how some patients resorted to quick-fix medicine to alleviate hypertension and diabetes symptoms, particularly consuming allopathic painkillers rather than or along with their prescribed medicine. Due to their daily household activities and daily wage work, patients focused on fast relief rather than comprehensive care. Low awareness regarding the symptoms of their disease often led patients to prefer quick-fix medications. For instance, one patient with hypertension told us:

> “*I just told them (the staff at the public health post) that I had pain in my legs and that my body was also paining. They gave me some medicine. I didn’t tell them that I have high BP (blood pressure). I just told them to give me some medicine for my body ache.*” (Female, diagnosed with hypertension more than 15 years prior to the interaction, age group 35 to 45 years)

The nature of residence and livelihood also influenced mixed medication patterns. For instance, some residents in the settlements studied were seasonal migrants, owing to the industrial/commercial job opportunities in the region-who managed their disease through a mix of medicines from their home town and their place of work-

> *“My doctor is in my home town (400km away). I get two to three months of BP(blood pressure) allopathic medicine from there when I come here. If I have any problem here, I go to the local doctor (unani). He writes me a different medicine and I take this pill (from the allopathic doctor) in the morning and his pill (from the unani doctor) in the evening.”* (Male, diagnosed 11 to 15 years prior to the interaction, age group more than 65 years)

##### Changing medications

Several patients reported changing their medication for reasons other than clinical recommendations. These changes often included alternating between multiple treatment regimens prescribed by different doctors they consulted. Patients reported following these treatment regimens according to their beliefs and convenience, often because of poor doctor-patient relations and low trust in doctors. For instance, in Case 2, the patient switched to a different treatment regimen for diabetes control since she believed that the previously prescribed medicine did not agree with her.

People also attempted to switch to cheaper medication and constantly explored the options available to them:

> “*The medical store suggested that I could try a similar medicine from another company at half price, so I took one strip. But it did not suit me, so I went back to my old medicines.*” (Female, diagnosed with hypertension 3 to 5 years prior to the interaction, age group 56 to 65 years)

These options were most often explored on the advice of neighbors or owners/workers of pharmacies and without formal doctor consultation.

##### Breaks and stops in medication

Around half the patients we spoke to reported taking breaks from their prescribed medications or stopping these completely. These breaks and stops often depended on patients’ perception of their disease severity. Several patients reported that they stopped medicines when they “felt better” and restarted them after symptoms resurfaced or worsened; Case 3 explains the travails of one such patient.

###### Case 3: Breaks in medication

A patient diagnosed with hypertension and diabetes two years prior to the interaction, took a break in his medicines when he felt better*-* “*I did not feel anything was wrong with me, so I stopped the medicines for a month.*” Thereafter, his eyes started hurting, and when he tried to soothe them, he got a cut in his eye that did not heal for two months. Following this, he went to a few eye specialists. One eye specialist related this issue to diabetes and enquired whether he had stopped any medicines-“*He asked me about diabetes, and I said I stopped the medicines of my own will. The doctor told me never to stop this medicine, that it would affect my eyes. After this incident, I decided I was never going to stop; I have it regularly now.*” (Patient, male, diagnosed upto 2 years prior to the interaction, age group 56 to 65 years)

We also encountered patients who felt compelled to stop medicines due to poor services at public health facilities. In one case, a patients’ poor experience during pregnancy in the public hospital and the rude attitude of the staff hindered her from going there for cheaper diabetes medicines:

> “*It was actually a problem with money, so I have stopped (medicines) for the last one and a half months. In the public hospital, if I ask something, they will answer back rudely. They have no consideration for us, we are also humans, but they never speak properly with us… whatever the problem is, we will not go to a public hospital. I will stop taking medicine for some time, but I will not go there.*” (Female, diagnosed with diabetes upto 2 years prior to the interaction, age group less than 35 years)

Further, we found that patients with comorbidities prioritized one medicine regimen over others. For instance, one patient stopped her diabetes medicine when she started treatment for tuberculosis:

> “*I stopped taking diabetes medicines on my own. I was taking tuberculosis medicines, so I stopped them. Should I have tuberculosis medicines or diabetes medicines? How many can I have?*” (Female, diagnosed with diabetes 11 to 15 years prior to the interaction, age group 46 to 55 years)

Comorbidities made it difficult to adhere to multiple treatment regimes, eventually leading to breaks in medicine regimens that were considered less problematic or urgent. When multiple treatments were prescribed, patients were inclined to prioritize one over the other at their discretion rather than seek medical advice.

#### 3.1.2 Lifestyle changes

Lifestyle changes are integral to comprehensive care for chronic diseases like hypertension and diabetes. In this section, we explore how patients tried to adhere to medical advice related to diet and exercise.

##### Partially following dietary advice

A majority of the patients reported partial adherence to diets as recommended by their healthcare providers (Refer to Case 4). The importance of diet in the comprehensive treatment of hypertension and diabetes was not always recognized by patients or shared by doctors, leading to patients partially following dietary recommendations as per their convenience and ability.

###### Case 4: Partially following dietary advice

A patient was diagnosed with diabetes during her pregnancy. After her delivery, she had stopped taking her diabetes injections and medicines. Later, when she felt unwell and did her tests, she found that her blood sugar had continued to remain high. So she restarted the medicines. She mentioned that she does not have any symptoms as of now. Her doctor has asked her to follow a diet, but she is able to do so only partially. Various reasons, such as attitudes and no symptoms, have led her to partially follow dietary prescriptions: “*It doesn’t happen suddenly (stopping certain foods like potatoes and rice), but yes, I try not to eat. I don’t have the habit of eating only roti (flatbread). Unless I eat rice, I don’t feel full. So, I must eat a little bit of rice.*” She further mentioned that there is no one to support her in the family. Therefore most of her disease management is based on as much as she can do for herself: “*They (doctors) say I should look after my diet properly, but I don’t even have anybody to look after me in the house.*” (Patient, female, diagnosed 6 to 10 years prior to the interaction, age group 35 to 45 years)

##### Following diets based on local knowledge

Few patients reported following dietary practices based on their perceptions and experiences of which diets were helpful:

> *“Sometimes in the night when I feel uneasy, I need to eat sweets say, chocolate, jaggery, biscuits etc. so I always have it on my bedside. The doctor has not told me this, I know this as I have high sugar for many years now.”* (Male, diagnosed with diabetes 11 to 15 years prior to the interaction, age group above 65 years)

Such diets were not prescribed or suggested by healthcare providers but often depended on advice from neighbors and family.

##### No dietary recommendations followed

Some patients reported that they were unable to follow diets or did not follow diets. Compulsions related to livelihood, finances, or household duties often prevented patients from adhering to diets even if they wanted to.

> *“The doctor has said to eat wisely, you can’t eat a lot of things. But I am unable to do it. I cook meals for around fifteen people from my village and earn some meager amount from that. So, I have to eat whatever I cook for others.”* (Female, diagnosed with diabetes 11 to 15 years prior to the interaction, age group 35 to 45 years)

Similar to the above quote, patients working in factories or warehouses had meals at their workplace, where the food was not customized as per their requirements, such as low salt or no sugar. For some patients, home-cooked meals were also limited to cereals and pulses; due to financial constraints, patients could not afford to have prescribed diets of fruits, vegetables, or eggs, which were perceived as more expensive.

At times, patients shared that the doctors they consulted had not given them any dietary advice. For instance, a hypertensive and diabetic patient does not follow any dietary restrictions: “*The doctor told me to eat what you want to; just don’t stop the medicine. So, I eat everything; I don’t limit eating any food*.” (Male, diagnosed upto 2 years prior to the interaction, age group 56 to 65 years)

##### No exercise recommendations followed

Except for a few patients, most did not specifically report exercising as a part of their medical advice. Exercise recommendations when received from doctors, were limited to suggestions for walking and doing physical activity, without providing a specific exercise regimen. Patients in our setting performed arduous work daily, both in factories and warehouses, and the household, which they considered exercise or physical activity. Doing physical activity in addition to their daily work was considered a remote possibility, as seen in Case 5.

###### Case 5: Daily work as exercise

A hypertensive patient mentioned that if she stops taking the medicine, she has severe body aches. The doctor has asked her to exercise and take walks, but she considers this a remote possibility given her situation: “*I am a housewife. Where can I go for a walk? I wake up early in the morning, prepare breakfast, clean the house and utensils, prepare food, and do other household things. Till I complete all my work, it’s three in the afternoon. I tell him (the doctor) that the housework gives me enough exercise.*” She recalls walking earlier to nearby places, but now she feels she is unable to walk due to her disease. (Patient, female, diagnosed 3 to 5 years prior to the interaction, age group 35 to 45 years)

Often covert in such narratives were the limitations arising from the context of urban informal settlements with dense and congested housing, inadequate access, and a lack of open spaces, which made “taking a walk” or exercising unfeasible. From the patient narratives, it appeared that most doctors did not prescribe exercise as a part of medical advice, highlighting a disconnect between lifestyle change recommendations that are otherwise an integral part of medical advice for chronic diseases, and the lived realities in our setting.

#### 3.1.3. Medical follow-ups

In this section, we explore how patients followed medical advice related to follow-up care to monitor their disease, such as visits to the doctor after diagnosis and routine testing for blood pressure and sugar.

##### Sporadic follow-ups

A majority of patients we spoke to underwent medical follow-ups only if they felt unwell or if their symptoms worsened. Most patients believed that follow-up visits to the doctor were not necessary and that they were not financially viable. Some patients went for repeat testing directly to pathology laboratories nearby and often self-interpreted the results without showing the reports to a doctor, as seen in Case 6.

###### Case 6: Sporadic follow-ups

A diabetic patient who tried several doctors shared that there were some doctors whose medicines did not suit and others who spoke rudely: “*Now, I don’t go to him (the doctor). I don’t go to any doctors now. I go to the lab on my own if I have to check.*” He takes medicines directly from the pharmacy. He does his blood sugar test based on convenience, “*if I feel like it.*” He directly goes to a nearby diagnostic laboratory to test his blood sugar without consulting the doctor. He only visits his local private family doctor if his sugar levels have increased ‘alarmingly’ in the test reports from his perspective. (Patient, male, diagnosed 3 to 5 years prior to the interaction, age group 46 to 55 years)

##### No follow-ups

Some patients shared that they did not go to the doctor for routine check-ups at all and/or did not do any follow-up tests to monitor their chronic conditions. Such patients continued their initially prescribed medications directly from the pharmacy.

> “*If at all I feel any discomfort, I buy medicine from the pharmacy and take it. If I go to the doctor to check my blood pressure, he will charge me fifty rupees. Instead, it is better to take the medicine.*” (Female, diagnosed with hypertension more than 15 years prior to the interaction, age group 35 to 45 years)

Financial constraints were reported as the major reason for not visiting doctors or doing regular tests, but our discussions also revealed that patients were often unaware of the need to monitor their disease regularly.

### 3.2 One patient, multiple ways of adherence

We have elaborated in detail on the various ways patients have attempted to adhere to medical advice in our setting. It is essential to note that each patient reported more than one way in which they tried to adhere to medical advice. Patients also shifted from ‘optimal’ adherence to ‘poor’ adherence and vice versa during their patient journeys. Therefore, the ‘poor’ adherence of each patient cannot be clubbed into one type or category and instead needs to be seen as a range of patient-wise adherence patterns, as represented in Table 2. To understand the adherence patterns holistically, Table 2 also includes ‘optimal’ adherence categories, i.e., those who had reported having adhered to medical advice on medication, lifestyle changes, and follow-ups. (Refer to Annexure Table 1A for details on patients who reported optimal adherence to medical advice).

**Table 2:**
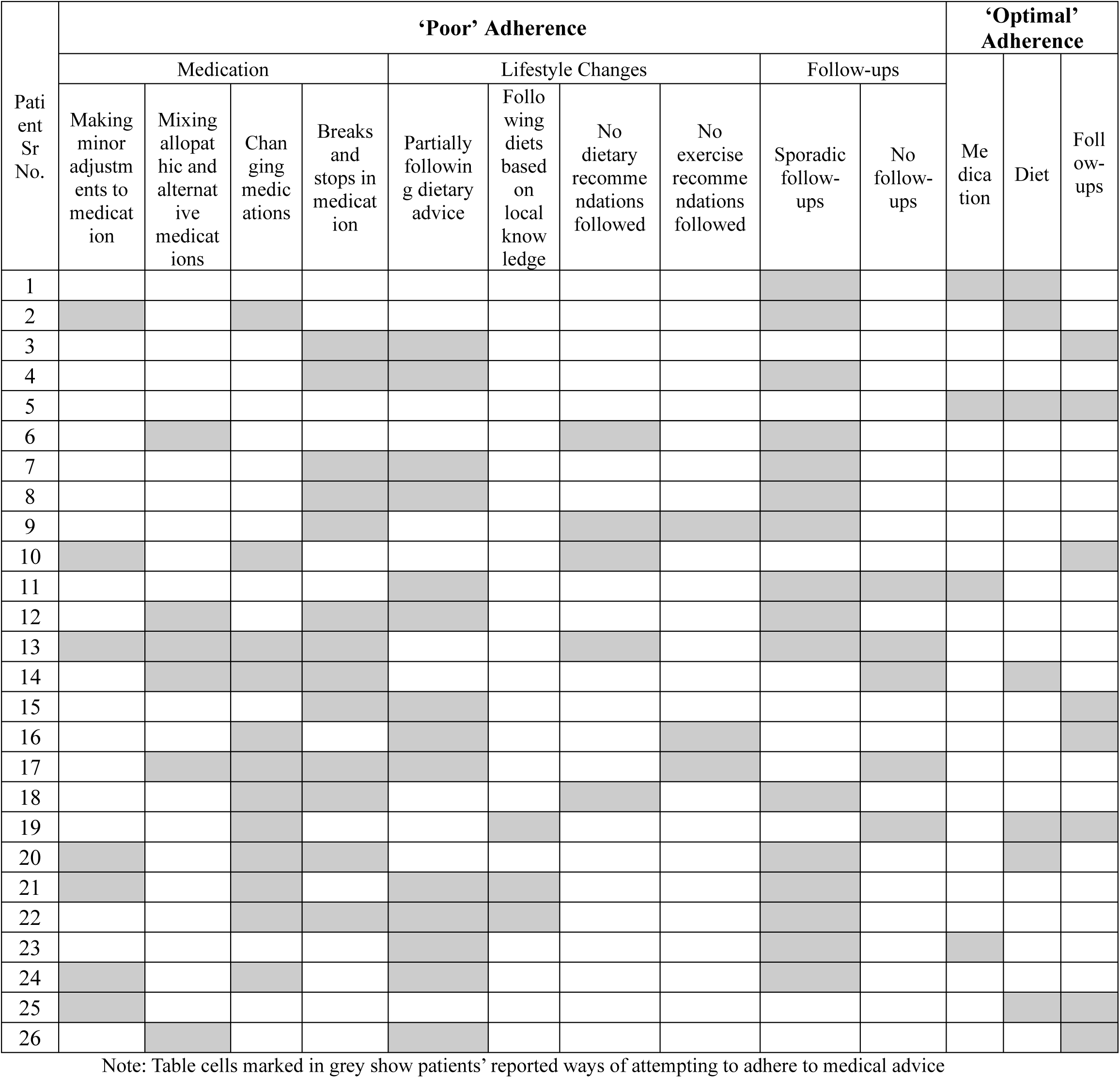
Patient-wise adherence patterns.

Table 2 shows that most patients were seen to adhere to medical advice partially. Various ways of changing medications and breaks and stops in medication were the most common types of ‘poor’ medication adherence. Partially following dietary measures and sporadic follow-ups characterized the most common adherence pattern among other medical advice.

Table 2 highlights the complexities faced in classifying the ways in which patients attempted to adhere to medical advice. Most patients reported more than one way in which they tried to adhere to medical advice, both simultaneously and over a period of time. For instance, in trying to adhere to medical advice, most patients reported some or all of the ways among making minor adjustments, mixing and changing medicines, and taking breaks or stopping medicines.

Many patients also reported optimal adherence in one category and poor adherence in another, showing that patients may adhere well to some aspects of their medical advice and not to others. For instance, some patients adhered well to dietary advice but not to medication.

Further, we also encountered some patients who showed shifts from optimal adherence to poor adherence within each category of medical advice. For instance, patients started with regular tests for their condition but stopped visiting doctors for any follow-ups over time.

## 4. Discussion

In this study, ‘poor’ adherence to medical advice has been discussed in three sub-categories of medical advice-medication, lifestyle changes and medical follow-ups. Within each sub-category we have discussed varied ways in which patients in our setting have adapted the advice given to them on the management of NCDs.

### 4.1 A typology of ‘poor’ adherence: What ‘poor’ adherence constitutes in our setting

One important contribution of this study is widening the present understandings of ‘poor’ adherence using empirical data from a resource-constrained LMIC setting. Based on these understandings, we have synthesized a typology for ‘poor’ adherence grounded in our data, as represented in Figure 1.

**Figure 1:**
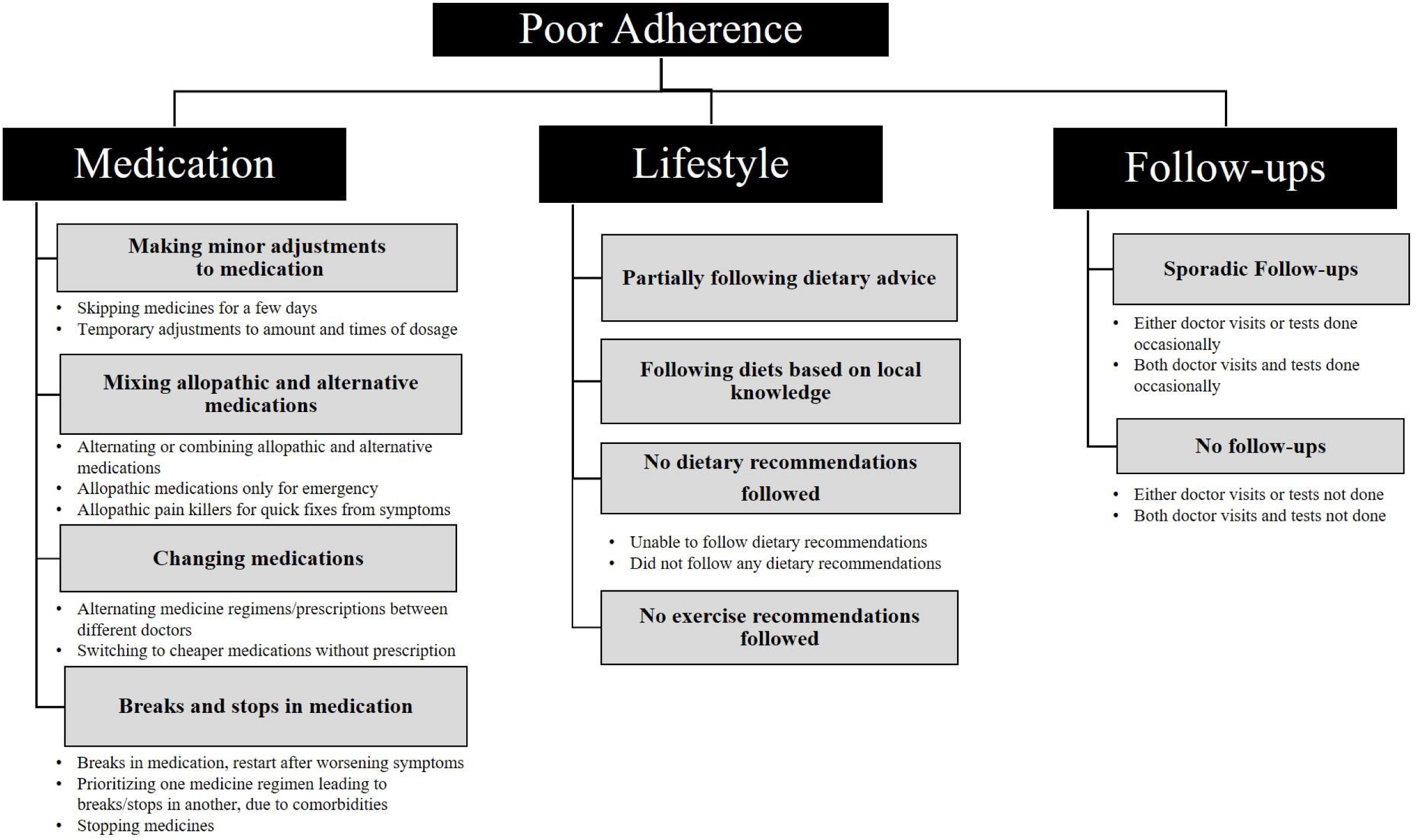
A typology of ‘poor’ adherence: What ‘poor’ adherence constitutes in our setting

This typology adds nuances to some of the existing typologies on adherence to medical advice. To begin with, it reemphasizes the need to think broadly about adherence to medical advice in the case of NCDs and shift focus from discussing only adherence to prescribed medication. This understanding is in agreement with the conceptualization of adherence proposed by other authors [39]. Since chronic diseases require, in addition to medication, lifestyle changes, and timely follow-ups, the conceptualization of adherence (or poor adherence) must also comprise all these components.

Our typology highlights that ‘poor’ adherence to medical advice in LMICs is a multifaceted concept. For instance, ‘poor’ adherence to medication can include varied practices such as adjusting dosages, mixing medicines, changing medicines, and taking breaks in medicine. Moreover, over time, patients traverse various categories of adherence presented in the typology, highlighting the complexity of conceptualizing ‘poor’ adherence in LMIC settings like ours.

Our typology is built from patient practices embedded in contextual specificities, making it more relevant to understanding adherence in vulnerable settings like ours. We found more commonly used typologies in existing literature, such as intentional and unintentional poor adherence [18,19], which separate the motivations and choice of patients leading to poor adherence (intentional) from the context and environment of the patient leading to poor adherence (unintentional), less helpful in our setting. In the urban informal settlements studied, patients’ choices and ‘intentions’ towards adherence were entwined with contextual limitations reported by them. For instance, familial support, particularly for male patients, enabled optimal dietary adherence, whereas financial constraints led to poor adherence to medication. Contextual specificities also shaped the diversity of adherence categories seen in our setting. Financial conditions and livelihood patterns in urban informal settlements, coupled with poor quality of public health services, lack of trust in local private doctors, and low education and awareness, led patients to adopt multiple ways to try to manage their diseases.

Similarly, other kinds of categorizations of adherence in literature-such as primary and secondary [20,22] offer linear understandings of poor adherence, which, too, do not fit well in explaining the convoluted care-seeking journeys experienced by patients in settings like ours [28,33,34]. Thus, our typology seeks to offer more grounded alternatives to conceptualizing ‘poor’ adherence that better explain LMIC contexts.

It was also challenging to characterize the term ‘medical advice’ in our study setting. First, medical advice received from doctors by patients in our setting was not always comprehensive-some patients reported not being prescribed a diet by their doctors or being provided with only temporary medicines for a few days. In these cases, the question of adhering to medical advice becomes moot since advice is not available to patients in the first place. Further, people obtained informal ‘medical advice’ not only from doctors but from multiple sources such as medicine sellers, neighbors and other patients. Adhering to this broad range of advice negates the very framing of adherence as ‘*concordance of medical advice between doctors and patients*’ [18]. All these complexities point to the need for developing more context-specific definitions of adherence. While existing typologies have been important in moving away from coercive definitions of adherence to being more cognizant of patient concerns [18, 19], our study points to the need to be cognizant of their limitations when applied universally. We also seek to problematize the use of the word ‘poor’ in ‘poor adherence’ – although this generally carries a negative connotation of deviation from prescribed medical advice, our study shows that ‘poor’ adherence is instead the multiple ways and practices through which patients continually juggle and try to adhere to medical advice, given the enablers and limitations of their context. We believe much more empirical work is required to frame and understand adherence to medical advice in LMIC settings.

### 4.2 Framing policy interventions based on the typology

The typology presented through our study can provide crucial inputs for framing better adherence strategies and policy interventions specific to patient needs in urban informal settlements.

Existing interventions are often based on reasons for poor adherence [3, 9] or universal categorizations such as intentional and unintentional adherence [8, 18]. Given the multifaceted nature of adherence in our context, our typology seeks to suggest health strategies that are better tailored to patient needs. This we explain through an example of health promotion strategies to improve adherence, which we as an organisation seek to adopt for future interventions in our setting.

Our typology shows that health promotion strategies cannot approach ‘poor adherence’ as a single uniform category but need to include more nuanced messaging based on patient practices and contextual influences identified. This includes specific messaging for medication, lifestyle changes and follow-ups, and the various sub-types of adherence within each category. Further, health promotion strategies need to be tailored based on contextual influences. For instance, patients who mixed and changed medications or took breaks due to financial constraints could be guided to access nearby generic stores and health posts where cheaper medicines are available. Since we found that patients often showed shifts from one form of adherence to another, multiple sets of messaging for various types and sub-types of adherence may need to be adopted based on identifying patients’ needs and practices related to adherence. Table 3 is our attempt at summarizing specific health messaging sets based on our typology.

**Table 3:** Health promotion messaging for patients based on our typology of ‘poor’ adherence.

| **‘Poor’ Adherence Typology** |  | **Health Promotion Messaging for Patients/their families on** |
| --- | --- | --- |
| Medication | Making minor adjustments to medication | • Importance of regular intake of prescribed dosage<br>• Adverse effects/worsening symptoms due to skipping medicines shown through other patient stories<br>• Not changing dosage amount and timings without consulting doctors<br>• Telling family members to remind patients to take medication regularly |
|  | Mixing allopathic and alternative medications | • Awareness of disease symptoms to avoid separate medications for symptomatic relief<br>• Need to inform doctors of other medicines taken previously/currently, including comorbidities<br>• Information about harmful effects of taking unprescribed/quick fix medicines<br>• Information on cheaper medicines available at generic stores/health posts/public hospital<br>• Encouragement to visit public healthcare facilities, reducing its negative perception through positive case stories |
|  | Changing medications | • Awareness of disease symptoms to avoid frequent changing of medication for symptomatic relief<br>• Need to inform doctors of other medicines taken previously/currently, including comorbidities<br>• Information on cheaper medicines available at generic stores/health posts/public hospital<br>• Encouragement to visit public healthcare facilities, reducing its negative perception through positive case stories<br>• Caution against buying medicines directly from pharmacy/other medicine sellers without consulting doctors |
|  | Breaks and stops in medication | • Information of adverse effects/worsening symptoms due to breaks/stops shown through other patient stories<br>• Need to continue medicines inspite of no symptoms<br>• Need to continue medicines inspite of comorbidities and other medicines taken for that<br>• Information on cheaper medicines available at generic stores/health posts/public hospital<br>• Encouragement to visit public healthcare facilities, reducing its negative perception through positive case stories |
| Lifestyle | Partially following dietary advice | • Importance of diet in controlling diseases<br>• Informing family members/food providers about healthy dietary options, healthy recipes for patients |
|  | Following diets based on local knowledge | • Importance of informing/consulting doctor about dietary habits and diet options followed |
|  | No dietary recommendations followed | • Importance of diet in controlling diseases<br>• Awareness about available healthy dietary options at a low price in the vicinity, healthy recipes for patients |
|  | No exercise recommendations followed | • Importance of being physically active, especially for patients not engaging in any physical activity or sitting/working in one position for prolonged periods<br>• Doing physical fitness activities such as breathing exercises, spot jogging, stretching etc. more possible in the given context<br>• Walking to and from home to work/marketplace/transport station |
| Follow-ups | Sporadic follow-ups | • Need for regular testing every 3 to 6 months, irrespective of symptoms<br>• Information of cheaper testing facilities at nearby public health posts/dispensaries, no need to go to secondary or tertiary centers<br>• Encouragement to visit public healthcare facilities, reducing its negative perception through positive case stories |
|  | No follow-ups | • Need for regular testing every 3 to 6 months, irrespective of symptoms<br>• Information of cheaper testing facilities at nearby public health posts/dispensaries, no need to go to secondary or tertiary centers<br>• Encouragement to visit public healthcare facilities, reducing its negative perception through positive case stories |

Such tailored messaging can be implemented through several strategies outlined in the literature to improve adherence in LMICs [12,40,41]. Community-level messaging through community health workers and peer support groups is one such strategy, which has shown success in LMIC contexts [41, 42, 43]. ‘Adherence clubs’ [44] in slum settlements of Nairobi, Kenya, for instance, have benefitted patients by following an integrated approach of providing medicines, health education, group support and timely follow-ups [45].

It is important to note that while we have provided an example of health promotion strategies, our typologies can also help identify other policy interventions needed to improve adherence. For instance, patients changing or stopping medicines due to financial constraints require along with information on cheaper generic medicines available, health system-related interventions such as training and sensitization of healthcare providers/pharmacies [46], and easy access and availability of affordable medicines [47] at public healthcare services. Similarly, for patients who could not follow diets due to livelihood-related factors, along with awareness, community initiatives such as diet kitchens providing hypertension and diabetes-friendly food can be incentivized [48,49].

While the study offers a nuanced typology for ‘poor’ adherence and insights into addressing the issue, it has some limitations. The information gathered from patients about their adherence to medical advice is based on memory recall and could have excluded certain details that patients did not share with us or did not remember due to comorbidities or long patient histories. Also, in the present paper, we have reflected on adherence only from the perspective of patients and not included the perspectives of healthcare providers towards adherence. Although we have diversified the participants, since the patients were sampled through SNEHA’s existing program in the community, the selection of participants included mainly families. We did not have access to recently migrated male workers who were single and living in this area, and we acknowledge that their adherence patterns might have been different.

In summary, our study shows how existing conceptualizations and categorizations of adherence can be deepened further with more empirical work from LMIC settings. Nuanced understandings of adherence based on grounded research can help frame better policy interventions to improve poor adherence to medical advice on NCDs. This study also constructs a framework that can be adapted further in research and practice settings.

## Supporting information

Annexure Table 1

## Data Availability

No quantitative datasets were used in this study. De-identified transcripts of the interviews with participants can be provided by the corresponding author upon reasonable request. The applicant must have clearance from their local ethics board before accessing this data.

## Acknowledgments

We express our gratitude to all the individuals who participated in this study, as their valuable perspectives and experiences made it possible. We extend our appreciation to the SNEHA team working in these urban informal settlements for their contributions and support. We acknowledge the field support provided by Clipsy Banji, Sujata Tambe, Kalyani Upadhayay and their team. Lastly, we would like to thank the members of the SNEHA Research Group for their valuable feedback.

**Supporting Information Caption: Table A1: ‘Optimal’ Adherence to Medical Advice**

