## Supplementary material for "What constitutes ‘poor’ adherence to medical advice for chronic diseases? Insights from a qualitative study among hypertension and diabetes patients in urban informal settlements, Mumbai Metropolitan Region": Annexure Table 1

***Table A1: ‘Optimal’ Adherence to Medical Advice***

| **Optimal Adherence** | **Details** | **Quotes** |
| --- | --- | --- |
| Optimal Adherence to Medication | Few patients reported adhering to prescribed medication.  Financial stability, support of family and advice from healthcare providers influenced adherence to medication. | “*If it is cough or cold and you don’t feel like having medicines is different, but this (hypertension and diabetes) I have to have. I have not stopped it (medicines), because if I stop and I get worse it will not be good. If I forget to take my medicines, my children remind me. They take care of me.*” (Female, diagnosed with hypertension and diabetes 3 to 5 years prior to the interaction, age group 46 to 55 years) |
| Optimal Adherence to Diet | Some patients mentioned adhering to dietary advice.  Awareness about the importance of diet, support from the family and advice from healthcare providers were major reasons for adherence.  Gender-based roles in the cultural context of our setting also led women to “take care” of other family members, particularly males in managing diets. | “*My wife helps in diet control. I eat only one spoon of rice, not like earlier times. She does not give me potato. I have two daughters-in-law; she has told them to not give me anything which can harm me. They don't give me.*” (Male, diagnosed with diabetes upto 2 years prior to the interaction, age group 46 to 55 years) |
| Optimal Adherence to Follow-ups | Some patients shared that they regularly visited doctors or did tests after starting their treatment.  Such regularity was reported mainly in cases where other family members were also required to visit the doctor/do check-ups or in cases where getting medicines was contingent on visiting doctors/doing tests. | “*Every two to three months I do the checkup at the nearby lab. My husband also has diabetes, so I go with him, and we both do the checkup.*” (Female, diagnosed with diabetes 11 to 15 years prior to the interaction, age group 35 to 45 years) |
